# A systematic review and meta-analysis of the immunogenicity and safety of Chikungunya vaccine

**DOI:** 10.1101/2025.09.17.25335965

**Authors:** Li Yutong, Jiang Feng, Zhao Danting, Wang Yanna, Liu Yizong, Zhang Rongguang, Liang Wenjuan, Lu Jiahai

## Abstract

**Background:** Since July 2025, there have been outbreaks of chikungunya fever in some parts of China, which has drawn widespread social attention. Vaccination is the most economical and effective means of preventing and controlling infectious diseases. Currently, there are two chikungunya fever vaccines approved for marketing worldwide, and there is still a lack of systematic review and analysis of the immunogenicity and safety of this vaccine.

**Objective:** Compare the levels in immunogenicity and safety of Chikungunya (CHIKV) vaccine.

**Methods:** Computerized searches were conducted in PubMed, EMBASE, Web of Science (WOS), Scopus, Cochrane Library and Clinicaltrials.gov for randomized controlled trials on the implementation of CHIKV vaccines in human populations. The search period was from the establishment of the databases to August 2025. Two researchers independently screened the literature, extracted the data and assessed the risk of bias in the included studies. Data analysis was performed using RevMan 5.4.1 software.

**Results:** In terms of immunogenicity, the results showed that there was a statistically significant difference in seroreponse rate:*RR*=12.22 (95%*CI*: 9.05∼16.51, *I*^2^=89%, *P* <0.00001), as well as GMR: *RR*=13.88 (95%*CI*: 4.12∼46.79, *I*^2^=0%, *P*<0.00001). In the meta-analysis of safety indicators, there was a statistically significant differences in any adverse reactions:*RR*=1.30(95%*CI*: 1.18∼1.42, *I*^2^=45%, *P*<0.00001). The meta-analysis results of local adverse reactions showed that there was a statistically significant differences in any local adverse reactions:*RR*=1.61 (95% *CI*:1.42∼1.82, *I*^2^=67%, *P* < 0.00001), as well as injection site pain: *RR*=1.83 (95% *CI*: 1.46∼2.28, *I*^2^=38%, *P* < 0.00001). The results showed that there were statistically significant difference in systemic solicited adverse events: *RR*=1.54(95%*CI*:1.44∼1.66, *I*^2^=87%, *P*<0.00001), as well as their common manifestations(including headache, fatigue, myalgia, arthralgia, nausea, fever).

**Conclusion:** CHIKV vaccine has demonstrated well immunogenicity. However, further research is still needed on the adverse events following vaccination.

**Author Summary:** Chikungunya fever, as a neglected tropical disease, experienced a relatively severe outbreak in the Guangdong region of China in 2025. Vaccination, as the most effective vaccination method, currently only has two approved vaccines globally. This study systematically reviewed the published RCTs results of different technical routes of chikungunya fever vaccines that have been on the market and are still in clinical trials, to understand, analyze and master the various immunogenicity and safety data of chikungunya fever vaccines. Study found that vaccination with the chikungunya fever vaccine was able to induce strong neutralizing antibodies and demonstrated an excellent seroreponse rate. However, in terms of safety, some significant adverse reactions were observed, especially in the overall adverse reaction rates such as headache, fatigue, muscle pain, fever, etc., as well as the local adverse reaction rates such as pain at the injection site. This inspires us to pay more attention to the safety of chikungunya fever vaccines in the future vaccine research and development process.

## 1. Introduction

Chikungunya virus(CHIKV) is an enveloped, single-stranded, positive sense RNA virus. The viral RNA is translated from the full-length genomic RNA or a subgenomic RNA as two polyproteins[1]; one encodes the four non-structural proteins (nsP1-4) to form a replication complex that synthesizes the genome, and the other encodes the structural proteins (capsid, 6K peptide, and E1, E2, and E3 envelope proteins)[2]. The envelope proteins are the dominant antibody targets of the host immune response with E1 conferring membrane fusion and E2 responsible for cell receptor (MXRA8) binding to target cells[3]. CHIKV is a member of the Semliki Forest virus antigenic complex that affords cross-reactive adaptive immunity to other emerging pathogenic alphaviruses including O’nyong nyong (ONNV), Mayaro (MAYV), Una (UNAV), and Ross River viruses (RRV)[4]. CHIKV was first described in 1952 after an outbreak in people in Tanzania[5–7]. CHIKV is an alphavirus (Togaviridae, Alphavirus chikungunya) that comprises four major genetic lineages (West African, East Central South African, Asian, and Indian Ocean Lineage). Despite this genetic diversity, CHIKV comprises a single serologic group[8].

Vaccines are effective means and methods for disease prevention and response. There are currently two approved CHIKV vaccines on the global market: a live attenuated vaccine (IXCHIQ)[9] and a virus-like particle vaccine (PXVX0317)[10]. Other technical routes of CHIKV vaccines, such as mRNA vaccines and viral vector vaccines, are also in the pre-market clinical trial stage.The study collected all publicly available randomized controlled trial research on CHIKV vaccines to explore their immunogenicity and safety levels.

## 2. Method

### 2.1. Search strategy

We adhered to the Preferred Reporting Items for Systematic Reviews and Meta-analyses (PRISMA) 2020 guidelines in this systematic review and meta-analysis. The Population, Intervention, Comparison, Outcomes, and Study (PICOS) framework guided the formulation of our clinical questions. Our search for relevant studies spanned from the establishment of the databases to August 14, 2025, across several databases: PubMed, EMBASE, Web of Science (WOS), Scopus, Cochrane Library and Clinicaltrials.gov.

### 2.2. Full search strategy

Through our search on PubMed, we employed both MeSH term search and free-text word search strategies.Utilizing the MeSH terms combination, we identified 27 results for:((“Chikungunya vaccine”[Mesh Terms]) OR (“Chikungunya virus vaccine”[Title/Abstract]) OR (“Chikungunya fever vaccine”[Title/Abstract]) OR (“IXCHIQ”[Title/Abstract]) OR (“VLA1553”[Title/Abstract])) AND ((randomized controlled trial[Publication Type] OR controlled clinical trial[Publication Type] OR randomized[Title/Abstract] OR randomised[Title/Abstract] OR placebo[Title/Abstract] OR drug therapy[MeSH Subheading] OR randomly[Title/Abstract] OR trial[Title/Abstract] OR groups[Title/Abstract]) NOT (animals[Mesh Terms] NOT humans[Mesh Terms])).

For our free-text word search, we used the following terms on Embase, Scopus, Web of Science (WOS), Cochrane Library and Clinicaltrials.gov., yielding 100, 22, 26, 34 and 21 results, respectively: ((“Chikungunya vaccine”) OR (“Chikungunya virus vaccine”) OR (“Chikungunya fever vaccine”) OR (IXCHIQ) OR (VLA1553)) AND ((randomized controlled trial OR controlled clinical trial OR RCT)).

All search results were imported into Covidence Extraction 2.0[11] Covidence was utilized to detect and remove duplicates. Two independent researchers systematically reviewed the titles and abstracts of the retrieved articles on Covidence, casting votes as ‘yes,’ ‘no,’ or ‘maybe,’ based on predefined inclusion and exclusion criteria. Articles were included in the review if both researchers voted ‘yes.’ A third researcher resolved any discrepancies between the researchers. Subsequently, the full texts of these selected articles were assessed similarly to confirm compliance with the inclusion criteria.

### 2.3. Inclusion criteria

(1) Papers or research reports publicly published in the database regarding the immunogenicity and safety of CHIKV vaccines; (2) Research type: Randomized Controlled Trial (RCTs); (3) Research Object: ①Able and willing to provide informed consent (and assent, as applicable) voluntarily signed by participant (and guardian, as applicable); ②Generally healthy, in the opinion of the investigator, based on medical history, physical examination, and screening laboratory assessments; (4) Intervention measures: The experimental group was injected with CHIKV fever vaccine; The control group was injected with a placebo; (5) Outcome indicators: Immunogenicity indicators include: serum response rate, Geometric Mean Ratio (GMR); Safety indicators include: total adverse reaction reporting rate; Reporting rate of local (injection site) adverse reactions (including pain, swelling and redness at the injection site); The reporting rate of systemic adverse reactions (including fever, fatigue, argyalgia, headache, etc.); The rate of serious occurrence reports.

### 2.4. Data extraction and risk of bias assessment of included studies

We excluded non-randomized clinical trials, observational studies, case series, reviews, commentaries, editorials, opinions, and case reports. Moreover, articles published in languages other than English, studies on nonhuman subjects were also excluded. Articles lacking outcome indicators or unable to extract basic data were also excluded.We extracted data on efficacy and safety of CHIKV vaccines by using Extraction 2.0 in the Covidence software. We assessed the risk of bias using the Cochrane Risk of Bias 2 for RCTs (RoB 2)[12]. ROB 2 contains five domains, while ROBINS-I contains seven. Both use standard signaling questions to provide structural judgment about the risk of bias[13].

### 2.5. Statistcal analysis

In the immunogenicity analysis, we conducted a meta-analysis of the Seroresponse rate and Geometric mean titer(GMT). Standard errors (*SE*) were calculated using log-transformed *GMT* ratios and 95% confidence intervals (*CI*). In addition, we also generated forest plots to display their pooled estimates.

For the analysis of adverse events(AEs), meta-analyses for binary outcomes were conducted to calculate risk ratios (*RR*), comparing the vaccine group and the placebo group.Forest plots were used to visualize pooled effect estimates, and funnel plots were generated to assess potential publication bias.

Meta-regression was performed to investigate the impact of the AE measurement time on the estimates.Funnel plots were used to determine publication bias. Furthermore, an influence analysis function based on the leave-one-out method was used to identify studies that caused heterogeneity. Sensitivity analyses were performed by excluding individual studies. A heterogenicity of 0 % to 40 % was considered as “might not be important,” 30 % to 60 % as “may represent moderate heterogeneity,” 50 % to 90 % as “may represent substantial heterogeneity”, and 75 % to 100 % as “considerable heterogeneity”[14]. All analyses were performed using RevMan 5.4.1.

## 3. Results

Of 230 records identified, 8 randomized controlled trials met the inclusion criteria (Fig. 1). Data extraction and risk-of-bias assessments were completed for all studies. Three trials evaluated IXCHIQ, two examined PXVX0317, and the remaining three investigated, respectively, the unlisted mRNA-based CHIKV vaccine mRNA-1388, the measles-vectored candidate MV-CHIK (still under development), and the unlisted live-attenuated vaccine TSI-GSD-218.

**Fig 1.**
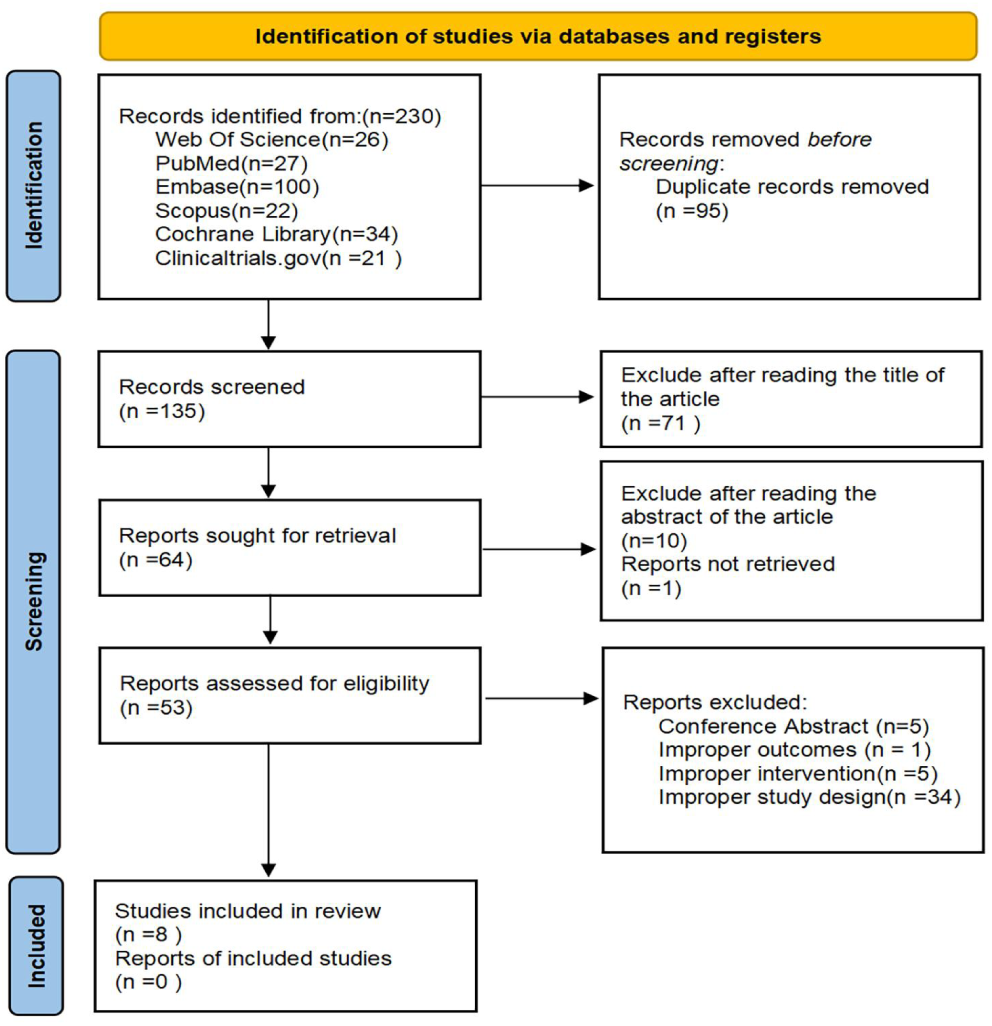
PRISMA flow diagram illustrating the selection process for studies on CHIKV vaccine.

All 8 articles reported information related to immunogenicity. Regarding the relevant indicators of safety, five articles reported Any adverse event, seven articles reported Any serious adverse event, any local solicited adverse event was reported in all studies except for Reisinger EC among the 8 articles, any systemic solicited adverse event was reported in all studies except for R Edelman. The characteristics of the studies included in the systematic review summarized in Table 1.

**Table 1.** Characteristics of the 8 studies included in the systematic review.

| First author, Year | Study design | Participants | Vaccine | Sample size | Female (n%) | Which information was reported? |
| --- | --- | --- | --- | --- | --- | --- |
| Jason SR, 2025[15] | Phase III, randomized, double-blind, placebo-controlled, parallel-group trial | Healthy adolescents and adults aged 12-64 years | IXCHIQ | 3,258 | 1,667 (51.2%) | Seroresponse rate, GMT; AEs; |
| Chen GL, 2020[16] | Randomized, placebo-controlled, double-blind, phase II clinical trial | Healthy adults aged 18 through 60 years | PXVX0317 | 400 | 199 (49.8%) | Seroresponse rate, GMT; AEs; |
| Lauren CT, 2025[17] | Phase III, randomized, placebo-controlled, double-blind, parallel-group, multicentre trial | Healthy participants aged 65 years and older | PXVX0317 | 413 | 242 (58.6%) | Seroresponse rate, GMT; AEs; |
| Edelman R, 2000[18] | Phase II, randomized, double-blind, placebo-controlled | Healthy volunteers, 18–40 years of age | TSI-GSD-218 | 73 | 26 (35.6%) | Seroresponse rate, GMT; AEs; |
| Reisinger EC, 2018[19] | Double-blind, randomized, placebo-controlled and active-controlled phase II trial | Healthy volunteers aged 18–55 years | MV-CHIK | 263 | 132 (50.2%) | Seroresponse rate, GMT; AEs; |
| Martina S, 2023[20] | Double-blind, multicentre, randomized, phase III trial | Healthy volunteers aged 18 years and older | IXCHIQ | 4,115 | 2,251 (54.7%) | Seroresponse rate, GMT; AEs; |
| Christine AS, 2023[21] | Phase I, first-in-human, randomized, placebo-controlled, dose-ranging study | Healthy adults aged $\geq 18$ years and $\leq 49$ years | mRNA-1388 | 60 | 34 (56.7%) | Seroresponse rate, GMT; AEs; |
| Vera B, 2025[22] | Double-blind, randomized, placebo-controlled phase III trial | Adolescents aged 12 to <18 years | IXCHIQ | 754 | 406 (53.8%) | Seroresponse rate, GMT; AEs; |

GMT, geometric mean titer; AEs, adverse events.

### 3.1 Immunogenicity

All eight included studies reported the Seroreponse Rate. The combined effect size *RR*=12.22 (95%*CI*: 9.05∼16.51, *I*^2^=89%, *P*<0.00001), indicating a statistically significant difference. The results demonstrated that the serum response rate in the vaccine group was significantly higher than that in the placebo group. The forest plot of Seroreponse Rate is shown in the following Fig 2(a).All eight included studies reported the GMT. The combined effect size *RR*=13.88 (95%*CI*: 4.12∼46.79, *I*^2^=0%, *P*<0.00001), indicating a statistically significant difference, the results indicated that the neutralizing antibody levels in the CHIKV vaccine group were significantly higher than those in the placebo group. The forest plot of GMR is shown in the following Fig 2(b).

**Fig 2.**
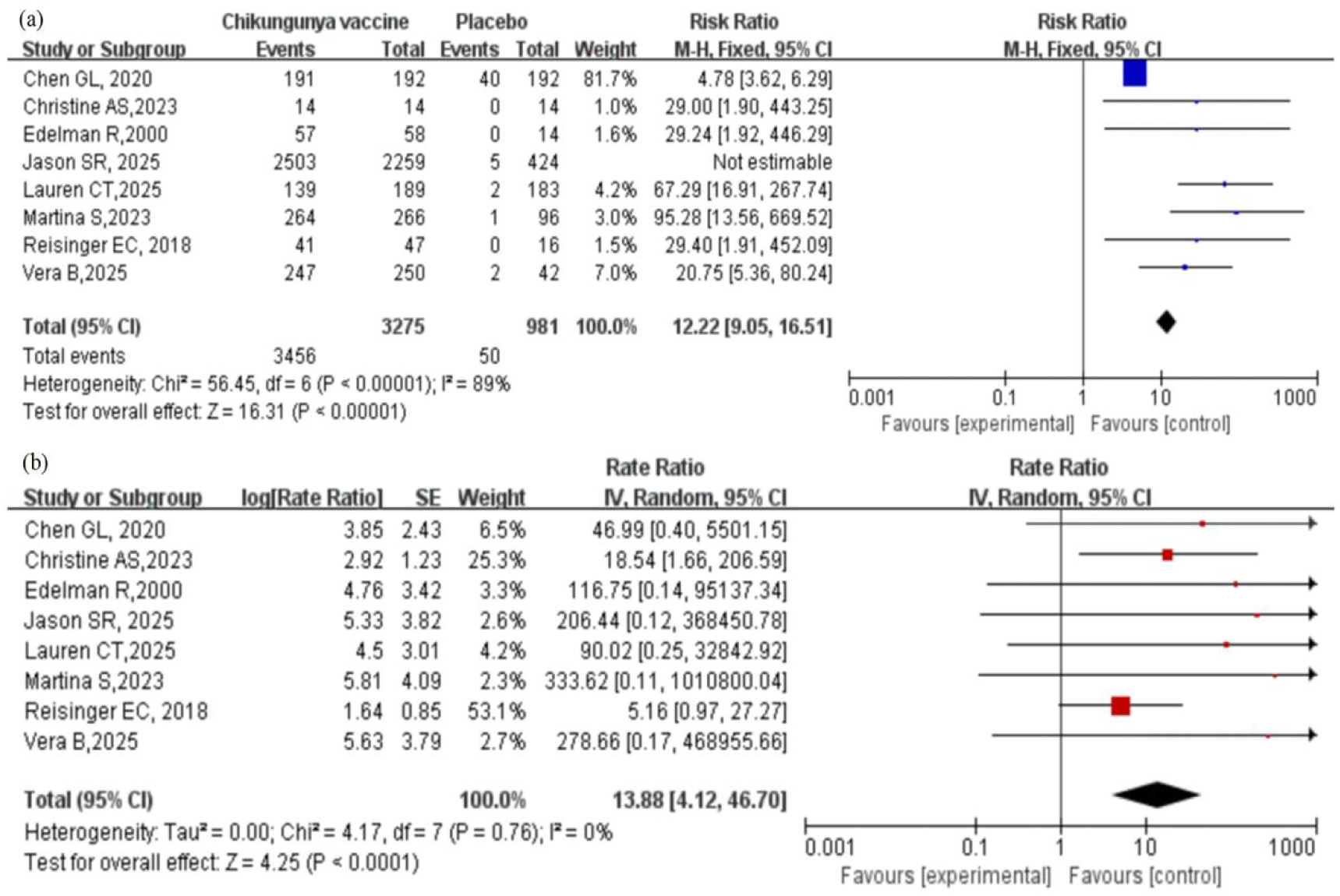
The forest plot of Immunogenicity for the CHIKV vaccine. (a) The meta-analysis results of the Seroreponse Rate between the CHIKV vaccine vaccination group and the control group showed a significant statistical difference: RR=12.22 (95%CI: 9.05-16.51, I^2^=89%, P < 0.00001). (b) The meta-analysis results of Geometric Mean Ratio between the CHIKV vaccine vaccination group and the control group showed a significant statistical difference: RR=13.88 (95%CI: 4.12-46.79, I^2^=0%, P < 0.00001)

### 3.2 Safety

Five articles were included among all the reported any adverse events, a total of 6,594 subjects were involved in the CHIKV vaccine group and 1,971 subjects in the placebo group, the results indicated that the risk of any adverse event in the CHIKV vaccine group was significantly higher than that in the placebo group, and the difference was statistically significant: *RR*=1.30(95%*CI*: 1.18∼1.42, *I*^2^=45%, *P*<0.00001). The forest plot of any adverse events is shown in Fig 3.

**Fig 3.**
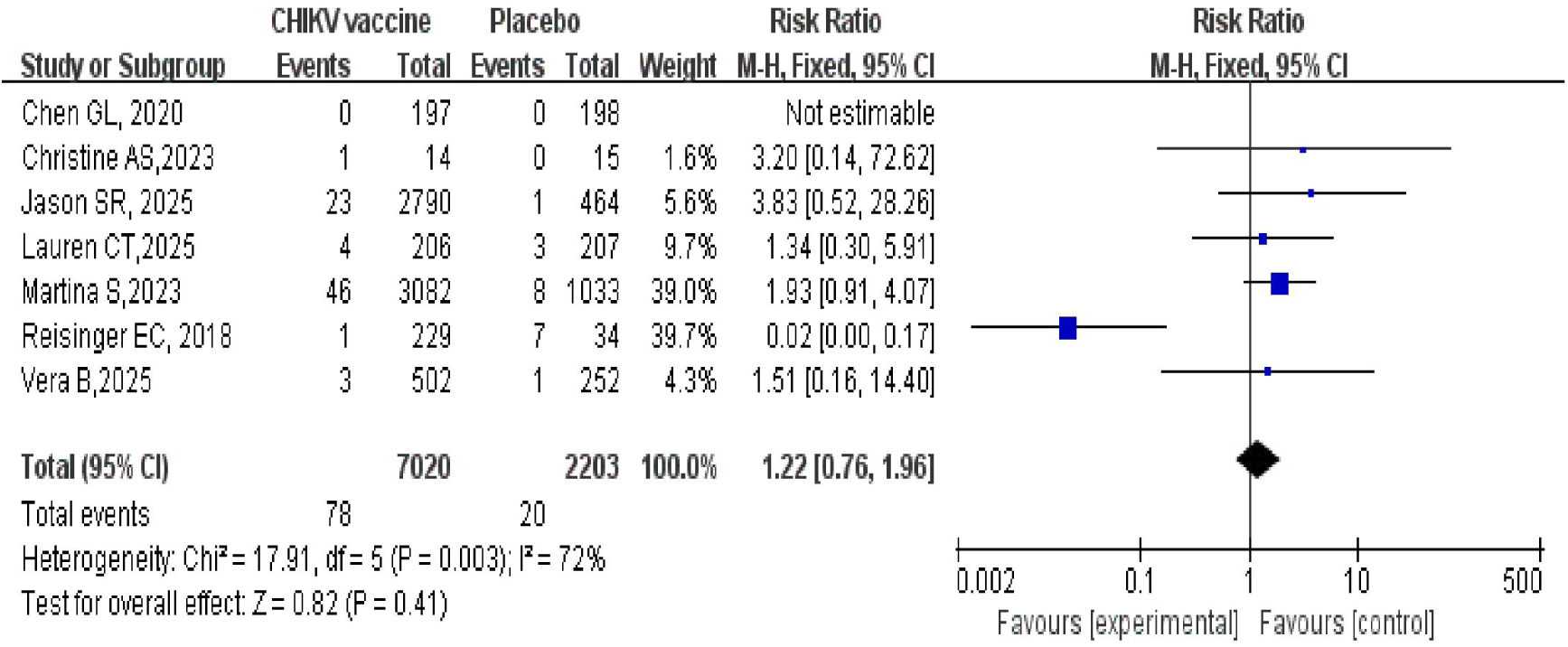
The forest plot presents the meta-analysis of SAEs reported for the CHIKV vaccine.

A total of 7 studies were included in this research, involving 7,020 subjects in the CHIKV vaccine group and 973 subjects in the placebo group, to assess the risk of serious adverse events (SAEs) of the vaccine. A fixed-effect model was used for the meta-analysis. The pooled results showed that there was no statistically significant difference in the risk of SAEs between the CHIKV vaccine group and the placebo group:*RR*=1.22(95%*CI*:0.76∼1.96, *I*^2^=72%, *P*=0.41). The forest plot is shown in Supplementary appendix.

#### 3.2.1 Local Solicited Adverse Event

Seven studies reported any local solicited adverse event. The pooled analysis showed that the risk of any local solicited adverse event in the CHIKV vaccine group was significantly higher than that in the placebo group, with a statistically significant difference: *RR*=1.61 (95% *CI*:1.42∼1.82, *I*^2^=67%, *P* < 0.00001). The forest plot of local solicited adverse events is shown in Fig 4(a).

**Fig 4.**
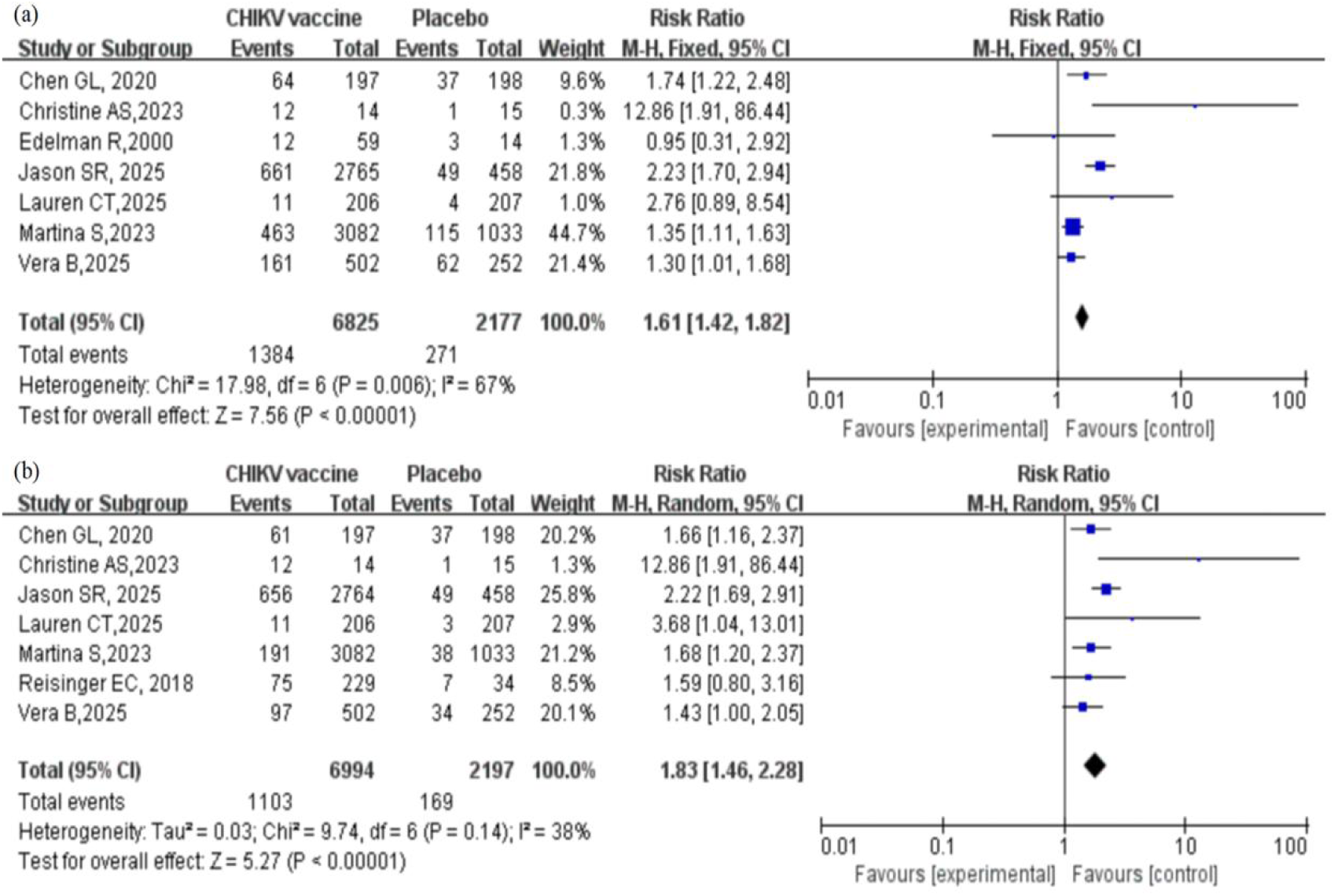
The forest plot of any local solicited adverse event and injection site pain. (a) Based on the meta-analysis results of Any Local Solicited Adverse Event reports, there was a statistically significant difference between the vaccination group and the control group: RR = 1.61 (95% CI: 1.42 - 1.82, I^2^ = 67%, P < 0.00001). (b) From the subgroup of Injection Site Pain, the difference between the vaccination group and the control group was also statistically significant: RR = 1.83 (95% CI: 1.46 - 2.28, I^2^ = 38%, P < 0.00001)

Injection site pain was the most common and prominent local reaction, with a total of 7 studies reporting on it. The results showed that the risk of injection site pain in the vaccine group was 1.83 times that of the placebo group, and the difference was statistically significant: *RR*=1.83 (95% *CI*: 1.46∼2.28, *I*^2^=38%, *P* < 0.00001). The forest plot is shown in Fig 4(b). Similarly, there were also 7 identical articles that reported on redness and swelling at the injection site, and the results showed that the differences were not statistically significant: *RR*=1.26 (95% *CI*:0.81∼1.96, *I*^2^=0%, *P*=0.31);*RR*=1.23 (95% *CI*: 0.50∼3.05, *I*^2^=38%, *P*=0.65). The forest plot of Injection site redness and swelling is shown in Supplementary appendix.

#### 3.2.2 Systemic Solicited Adverse Event

Seven articles reported systemic solicited adverse events, and the results showed a statistically significant difference, indicating that the risk of systemic adverse events in the vaccination group was higher than that in the placebo group: *RR*=1.54(95%*CI*:1.44∼1.66, *I*^2^=87%, *P*<0.00001). The forest plot of systemic solicited adverse events is shown in Fig 5(a).

**Fig 5.**
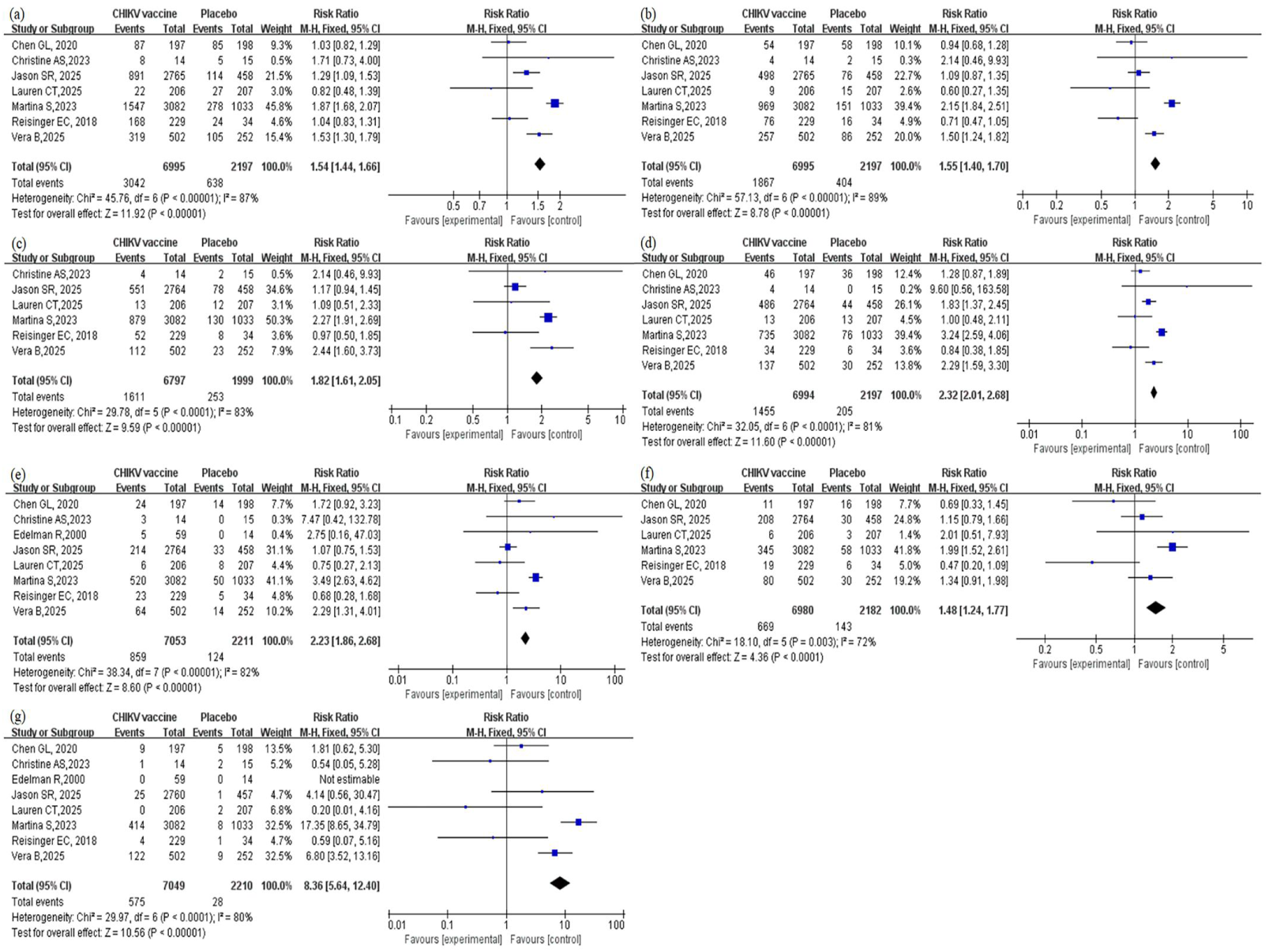
The forest plot presents the meta-analysis of systemic solicited adverse event and their Specific manifestations. (a) Based on the results of the meta-analysis of Systemic Solicited Adverse Events, there was a statistically significant difference between the vaccination group and the control group: RR=1.54(95%CI:1.44∼1.66, I^2^=87%, P<0.00001) (b) From the Headache subgroup, there was a statistically significant difference between the vaccination group and the control group: RR=1.55(95%CI: 1.40∼1.70, I2=89%, P<0.00001). (c) From the Fatigue subgroup, there was a statistically significant difference between the vaccination group and the control group: RR=1.82(95%CI: 1.61∼2.05, I2=83%, P<0.00001). (d) From the Myalgia subgroup, there was a statistically significant difference between the vaccination group and the control group: RR=2.32(95%CI: 2.01∼2.68, I2=81%, P<0.00001). (e) From the Arthralgia subgroup, there was a statistically significant difference between the vaccination group and the control group: RR=2.23(95%CI: 1.86∼2.68, I2=82%, P<0.00001). (f) From the Nausea subgroup, there was a statistically significant difference between the vaccination group and the control group: RR=8.36(95%CI: 5.64∼12.40, I2=80%, P<0.00001). (g) From the Fever subgroup, there was a statistically significant difference between the vaccination group and the control group: RR=8.36(95%CI: 5.64∼12.40, I2=80%, P<0.00001).

The results showed that in the reports of systemic solicited adverse events, the risk of all adverse events occurring in the vaccine group was higher than that in the placebo group. Headache was the most common adverse event:*RR*=1.55(95%*CI*: 1.40∼1.70, *I^2^*=89%, *P*<0.00001), the forest plot is shown in Fig 5(b); A total of 6 articles reported fatigue:*RR*=1.82(95%*CI*: 1.61∼2.05, *I^2^*=83%, *P*<0.00001), the forest plot is shown in Fig 5(c); A total of 7 articles reported on myalgia:*RR*=2.32(95%*CI*: 2.01∼2.68, *I^2^*=81%, *P*<0.00001), the forest plot is shown in Fig 5(d); A total of 8 articles reported on arthralgia:*RR*=2.23(95%*CI*: 1.86∼2.68, *I^2^*=82%, *P*<0.00001), the forest plot is shown in Fig 5(e); A total of 6 articles reported on nausea:*RR*=1.48(95%*CI*: 1.24∼1.77, *I^2^*=72%, *P*<0.0001), the forest plot is shown in Fig 5(f); A total of 8 articles reported on fever:*RR*=8.36(95%*CI*: 5.64∼12.40, *I^2^*=80%, *P*<0.00001), the forest plot is shown in Fig 5(g).

### 3.3 Risk of bias

All the 8 included studies mentioned the word "randomised", among which 4 used random number tables/codes, 3 studies adopted electronic systems such as interactive response system, and R Edelman’s study did not mention the random method. Regarding blinding, seven studies employed double-blind methods, and Vera Buerger’s research employed observer-blinded. All included studies reported allocation concealment and all outcome measures. Other sources of bias were unclear. The specific risk of bias assessment results are shown in Fig 6 and 7.

**Fig 6.**
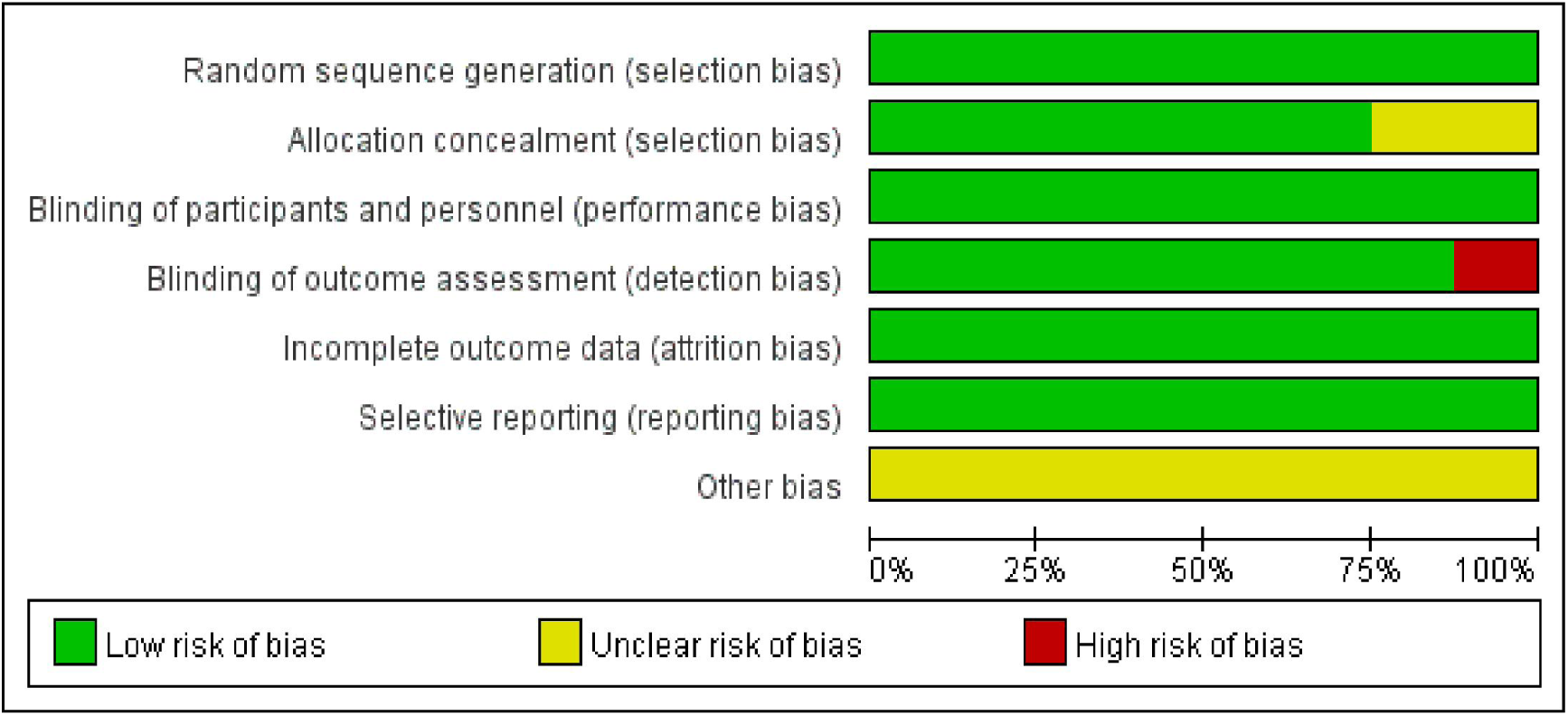
Risk of bias graph of included studies.

**Fig 7.**
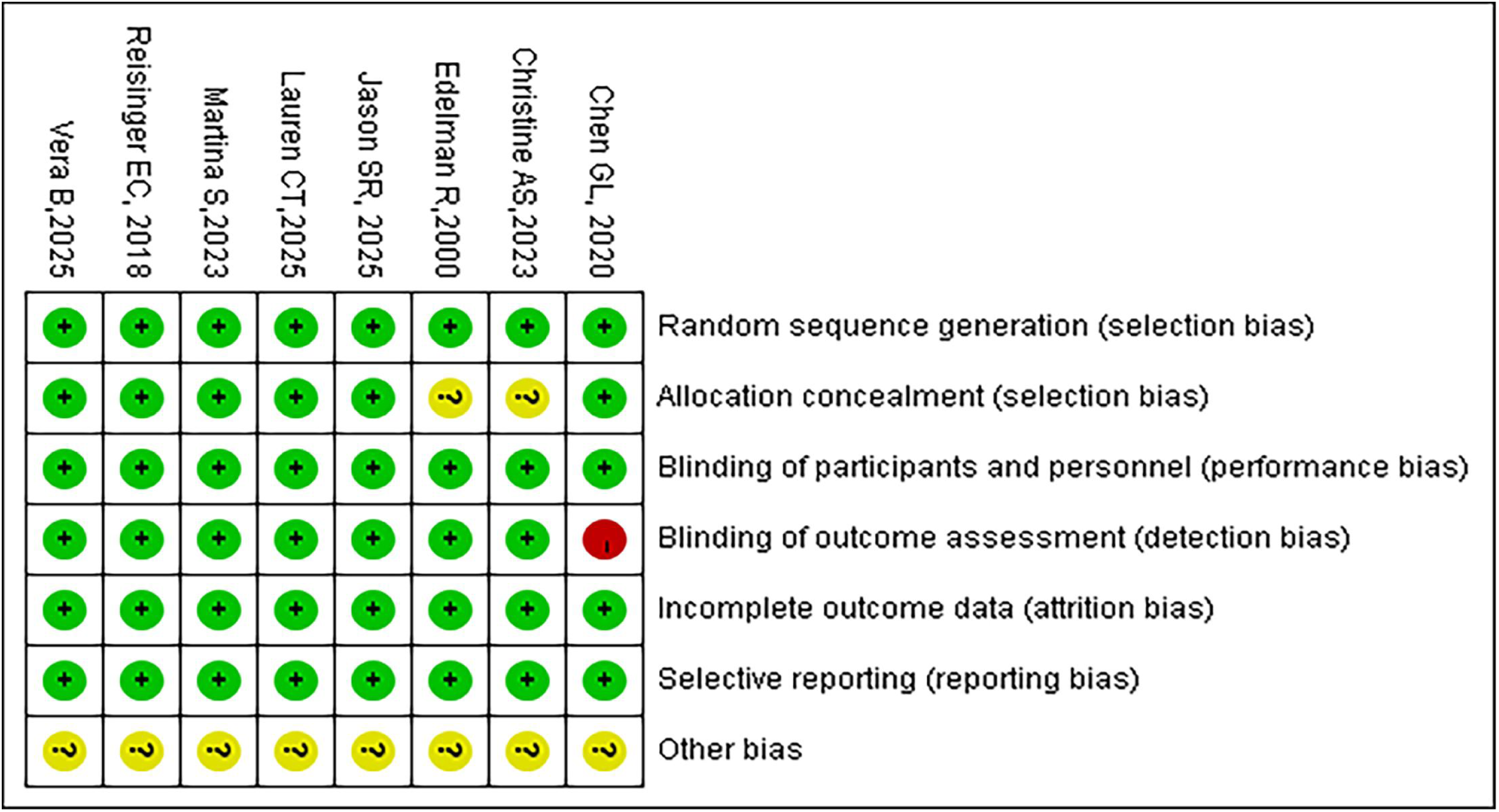
Risk of bias assessment graph for included RCTs.

## 4. Discussion

In this systematic review and meta-analysis, we mainly investigated the immunogenicity and safety of CHIKV vaccines. In terms of immunogenicity, there has been sufficient research and evidence to prove that vaccination with CHIKV vaccine can induces a rapid and robust immune response. The results of the meta-analysis of the serum response rate and neutralizing GMR values both demonstrated good protective efficacy. This is consistent with the research findings of Huang Zongyuan[23] and is also in line with the animal experiment results of Rossi et al[24]. and Saraswat et al[25].

In terms of safety, the results of the meta-analysis of various adverse reaction indicators show that the risk of occurrence of Any advance events, local solicited adverse event (including its specific adverse reactions: injected site pain), and systemic solicited adverse events (including its specific adverse reactions: headache, fatigue, myalgia, arthralgia, nausea, fever) are all reported to be higher than those in the placebo group. Notably, on August 22, 2025, the U.S. Food and Drug Administration (FDA) announced the suspension of the marketing authorization of Valneva’s CHIKV vaccine Ixchiq, citing "serious safety concerns"[26]. The statement pointed out that the Ixchiq vaccine "seems to cause patients who have been vaccinated to develop symptoms similar to CHIKV". The statement shows that up to now, the regulatory agency has recorded multiple reports of serious side effects in vaccinated individuals that are consistent with chikungunya-like symptoms. Although the FDA emphasized that the causal relationship between adverse events reported in VAERS and vaccination remains to be confirmed, the cautious attitude of the regulatory agency is clearly visible. After receiving the attenuated live vaccine (Ixchiq®), symptoms similar to mild viral infection (such as fever and joint pain) may occur, which could be due to the reversion of the attenuated virus to a more virulent form[27–29]. This is more risky for the elderly and those with immune deficiencies[30] After receiving the virus-like particle (VLP) vaccine (PXVX0317), adjuvant reactions, injection site reactions, and systemic reactions caused by the immune response (such as fever and fatigue) may occur[31–34]. After receiving the mRNA-based CHIKV vaccine (mRNA-1388), inflammatory reactions (such as fever and headache) may occur due to its lipid nanoparticles[35–37]. After receiving the viral vector vaccine (MV-CHIK), pre-existing immunity may affect its efficacy, and mild reactions related to the vector virus may occur[38–39]. However, the adverse reactions mentioned above are usually mild and self-limiting, which are normal manifestations of the immune system being activated. Studies have shown that there is no evidence to prove that the serious adverse reactions reported are related to the vaccination[40].

The risk of bias assessments indicated variability across studies, with some studies exhibiting concerns about randomization and selective reporting. This underscores the importance of careful study design and robust reporting practices in future trials to minimize bias and ensure the reliability of results.

The studies included in this review have several limitations, which make comparisons challenging. These limitations include variations in the methods used to analyze and report immunogenicity, the use of different vaccines, and discrepancies in participant numbers.

## 5. Conclusion

The ideal CHIKV vaccine should possess features such as single-dose protection, thermal stability and low cost. The research directions include optimizing the delivery system (such as developing freeze-dried formulations), exploring new adjuvants, and developing multivalent vaccines against co-endemic viruses (such as dengue fever and Zika virus).Our research results support that this vaccine has well immunogenicity, but its safety still requires long-term monitoring and further investigation.

## Data Availability

All data produced in the present work are contained in the manuscript.

## Funding

This work was funded by the National Natural Science Foundation of China International (Regional) Cooperation and Exchange Program, project number(2023YFVA1005), author: LJH, website: https://www.nsfc.gov.cn/;

National Natural Science Foundation of China, project number(82160634 and 72361127506), authors: ZRG & LJH, website: https://www.nsfc.gov.cn/;

the Education Department of Hainan Province, project number(Hnjg2025ZD-37), author:LWJ, website: http://edu.hainan.gov.cn/;

2024 Hainan Province Postgraduate Innovation and Entrepreneurship Project, project number(Qhys2024-424), author:LYT, website: http://edu.hainan.gov.cn/.

## Author Contributions

Conceptualization, Jiahai Lu, Wenjuan Liang and Yutong Li; Funding Acquisition, Jiahai Lu; Rongguang Zhang, Yutong Li; Investigation, Yutong Li, Feng Jiang; Methodology, Danting Zhao, Yanna Wang and Yizong Liu; Writing—Original Draft Preparation, Yutong Li, Feng Jiang; Writing—Review & Editing, Rongguang Zhang, Jiahai Lu and Wenjuan Liang. All authors have read and agreed to the published version of the manuscript.

**PROSPERO ID:** CRD420251091292

## Conflict of interest

All authors declare no conflict of interest.

## Notes

### Competing Interest Statement

The authors have declared no competing interest.

### Funding Statement

This study was funded by the National Natural Science Foundation of China International (Regional) Cooperation and Exchange Program, project number(2023YFVA1005), author: LJH, website: https://www.nsfc.gov.cn/;
National Natural Science Foundation of China, project number(82160634 and 72361127506), authors: ZRG & LJH, website: https://www.nsfc.gov.cn/;
the Education Department of Hainan Province，project number(Hnjg2025ZD-37), author:LWJ, website: http://edu.hainan.gov.cn/;
2024 Hainan Province Postgraduate Innovation and Entrepreneurship Project, project number(Qhys2024-424), author: LYT, website: http://edu.hainan.gov.cn/.

